# Genome wide association study of vaginal microbiota genetic diversity in French women

**DOI:** 10.1101/2025.05.02.25326864

**Authors:** Samuel Alizon, Claire Bernat, Vanina Boué, Sophie Grasset, Soraya Groc, Tsuksuhi Kamiya, Massilva Rahmoun, Christian Selinger, Nicolas Tessandier, Marine Bonneau, Vincent Foulongne, Christelle Graf, Jacques Reynes, Michel Segondy, Vincent Tribout, Jacques Ravel, Nathalie Boulle, Carmen Lia Murall, Vincent Pedergnana, Jean-François Deleuze

## Abstract

The composition of the vaginal microbiota is known to be highly structured into five main community state types (CSTs) that are found in all human populations. Several associations between self-reported ethnicity and the type of community have been reported. Analysing data from in 168 women from the PAPCLEAR cohort study in France, we perform a genome wide association studies (GWAS) looking for human genetic polymorphisms that may impact vaginal microbiota community composition. We show that Shannon diversity is the trait related to the vaginal microbiota that is best explained by the human genome. Furthermore, we identify two genomic regions associated with its variations. This is one of the first GWAS to use microbial genetic data instead of symptoms to characterise the vaginal microbiota and our results call for more powered studies in terms of participants and genome coverage.

## Introduction

The vaginal microbiota has been studied since the end of the 19th century, which is consistent with the massive impact it is now known to have on women’s susceptibility to sexually-transmitted infections (van de Wijgert, 2017), fertility (Haahr et al., 2019), and general well-being (Bilardi et al., 2013). The vaginal microbiota is also highly structured into five main community state types, or CSTs (Ravel et al., 2011), which tend to be relatively stable over several months (Tamarelle et al., 2024; Kamiya et al., 2025). Its composition has also been long reported to be associated with perceived ethnicity by several studies (Zhou et al., 2007; Ravel et al., 2011; Fettweis et al., 2014; Borgdorff et al., 2017). However, contrarily to other human microbiota, few studies have attempted to identify potential genetic factors associated with the observed variation in human vaginal microbiota (Kurilshikov et al., 2021).

A recent exception comes from a Genome Wide Association Study (GWAS) study in the USA on 686 women living with and at risk for HIV infection, the trait of interest being the presence or absence of bacterial vaginosis (BV) (Murphy et al., 2024). Another exception comes from a study in Kenya that performed a GWAS on 171 women (Mehta et al., 2020), using as a response trait the presence or absence of one of three bacterial species, the Shannon diversity, or the CST. Another approach also used bacterial 16S data to perform a GWAS with the presence of a variety for 359 pregnant women in China (Fan et al., 2021), one limitation of this study being that pregnancy is known to affect vaginal microbiota composition. Finally, a recent study performed a GWAS on 12,815 women living in Estonia and identified single nucleotide polymorphisms (SNPs) associated with recurrent vaginitis (Mutli et al., 2024).

Few studies investigated the association between the vaginal microbiota community and human genomics. This is surprising since all studies conducted around the globe have identified the same five main community state types (CSTs). Four of these communities are dominated, sometimes exclusively, by a single species of Lactobacillus (*L. crispatus* for CST-1, *L. jenseni* for CST-2, *L. iners* for CST-3, and *L. gasseri* for CST-5). Conversely, the fifth, CST-4, is a diverse assemblage of anaerobic bacteria from genera such as *Gardnerella*, *Prevotella*, or *Fannyhessea*. This latter CST is often more associated with BV and other health issues.

The relative proportion of each CST can vary widely between populations. CST-1, CST-3, and CST-4 are always the most frequent. However, in North America, CST-4 is four times more common in women who identify themselves as ‘black’ or ‘hispanic’, than in those who identify themselves as ‘white’ (Ravel et al., 2011). These differences could result from behavioural differences. For example, vaginal douching is known to be strongly associated with CST-4 and this practice varies across populations (Brotman et al., 2008). However, to date, we do not know to what extent this difference is associated with human genetics.

One of the motivations to identify potential SNPs associated with CST-4, also sometimes referred to as ‘molecular bacterial vaginosis’ (McKinnon et al., 2019), is that half of the time they are not associated with symptoms. Furthermore, long-term longitudinal follow-ups reveal that some women remain in CST-4 whereas others only occasionally shift to this CST (Tamarelle et al., 2024; Kamiya et al., 2025). One hypothesis is that some women can better tolerate these CSTs than others.

In this study, we performed a GWAS on 168 women from the PAPCLEAR cohort, which was designed to analyse human papillomavirus infections (Tessandier et al., 2025), using longitudinal vaginal microbiota metabarcoding data (16S) as a response trait. This work is of limited statistical power but stands out in terms of the traits considered and of the study population.

## Results

### Cohort description

The PAPCLEAR study enrolled women aged from 18 to 25 years old, living in the area of Montpellier (France), and primarily university students (119/138, 86 %). The main characteristics of the study population have been reported in earlier studies and some are shown in Table 1 for the samples analysed here and stratified in terms of vaginal microbiota composition. Most of the associations observed are expected based on earlier studies since women with a CST4, i.e. few lactobacilli, report lower condom usage from the male partner, higher lifetime number of sexual partners, and identify themselves less as ‘caucasian’ (Ma et al., 2012). Their body mass index (BMI) and age are also higher, and they have more often been pregnant. In terms of CST differences, the most frequent is CST-1, followed by CST-3, and then CST-4. As expected, Shannon diversity is much lower in the lactobacillus-dominated microbiotas (Ravel et al., 2011).

**Table 1:**
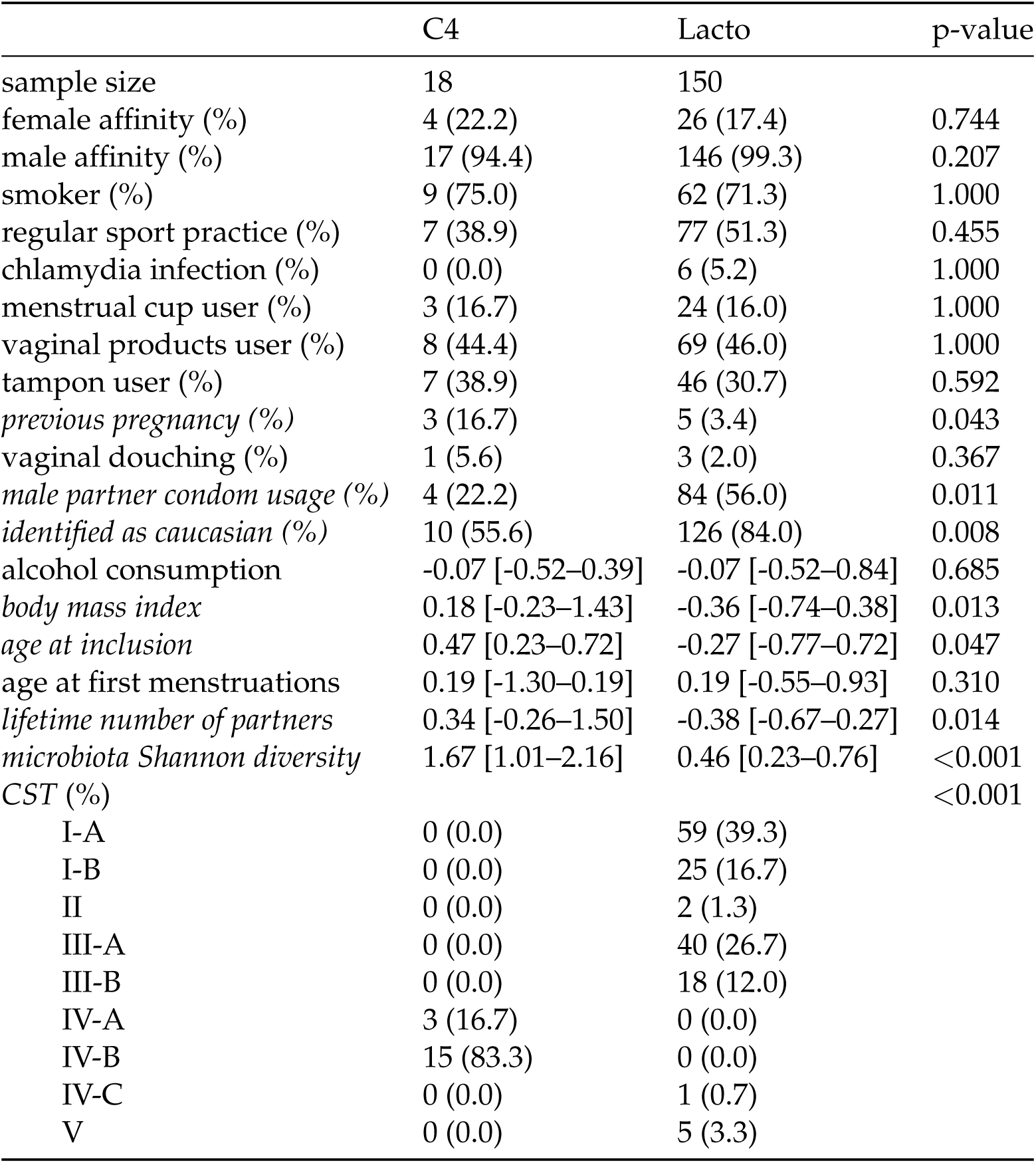
Cohort profile stratified by vaginal microbiota composition. Numerical variables are centred and scaled. The p-value indicated the outcome of a Fisher exact test for categorical variables and a Kruskal-Wallis rank sum test for continuous variables (the interquartile range is shown in brackets). Significant differences are in italic.

As expected, the genetic composition of the large majority of the study population corresponds to the individuals from European ancestry in the 1000 genomes database (Figure 1). Given the limited size of the sample, we chose not to remove the participants far away from the European-ancestry cluster but used the MDS coordinates as covariates in the association study as a control.

**Figure 1:**
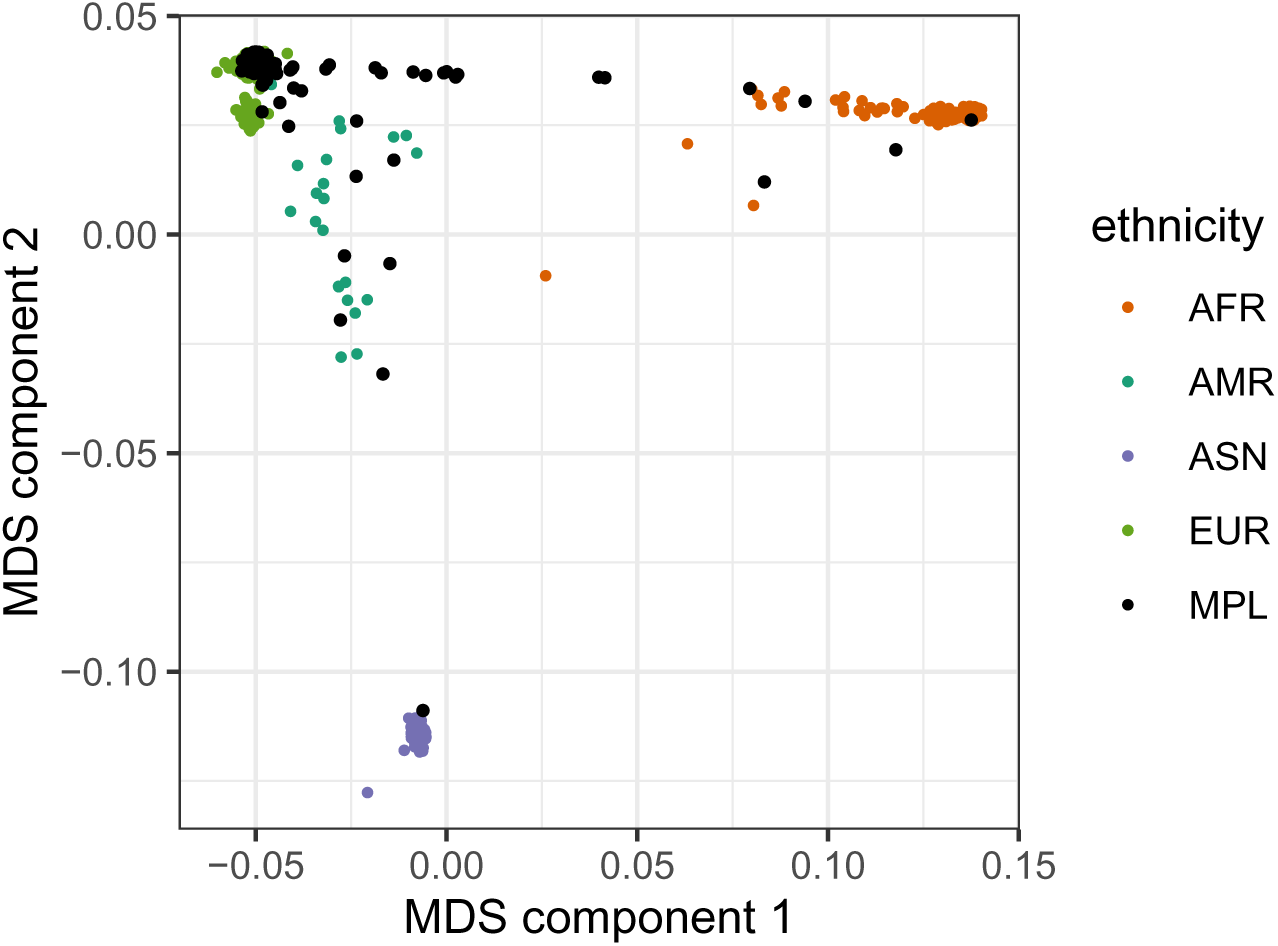
Multidimensional scaling (MDS) plot of population stratification. The four colours show the main ancestries for the 1000 Genomes database. The PAPCLEAR study participants from Montpellier (‘MPL’) are shown in black.

**Figure 2:**
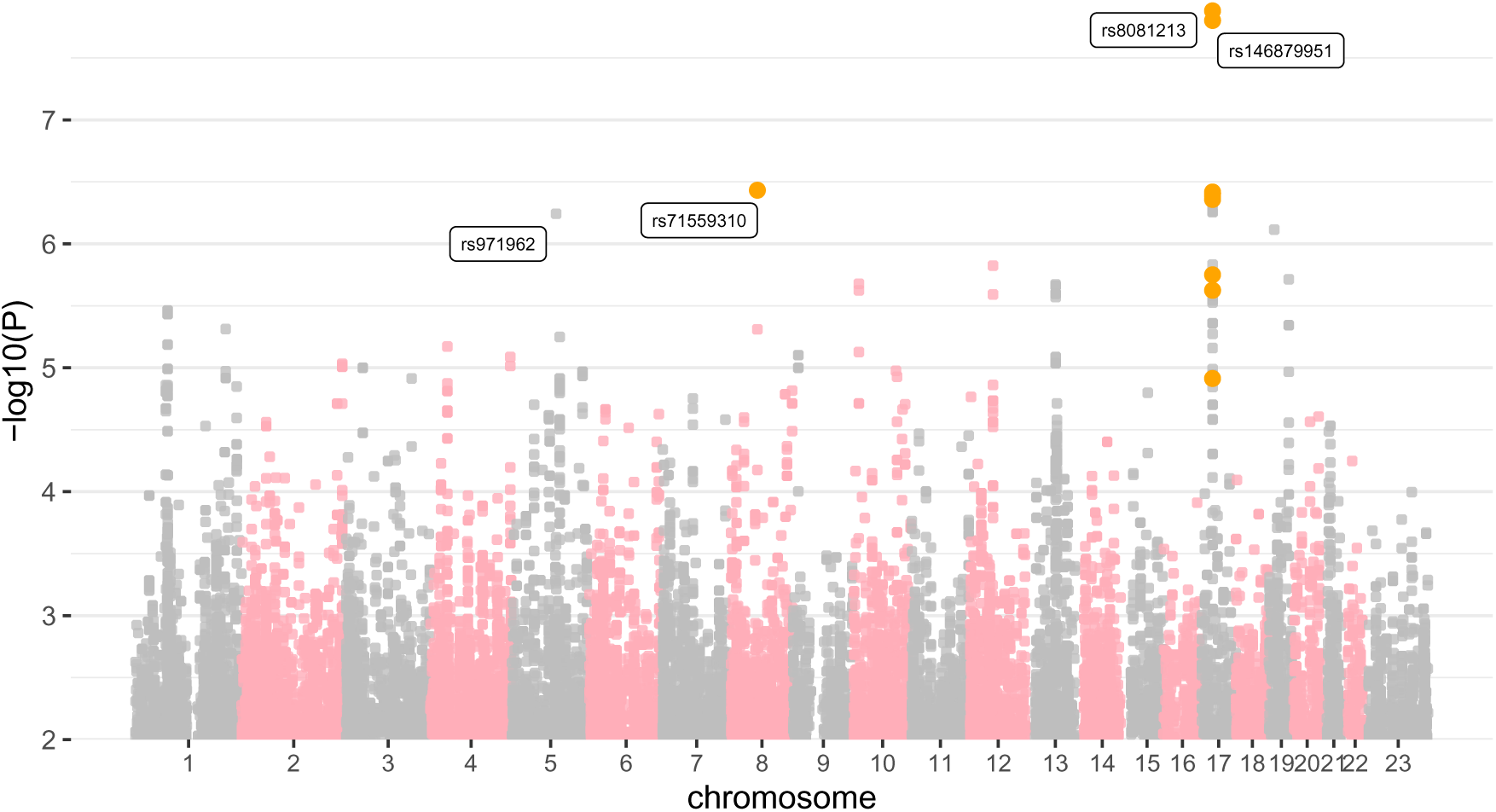
Manhattan plot showing associations with vaginal microbiota Shannon diversity. Ancestry is taken into account in the regression via the MDS components. Colours indicate SNPs or regions of interest, which are also labelled. The X chromosome is labelled as ‘23’.

**Figure 3:**
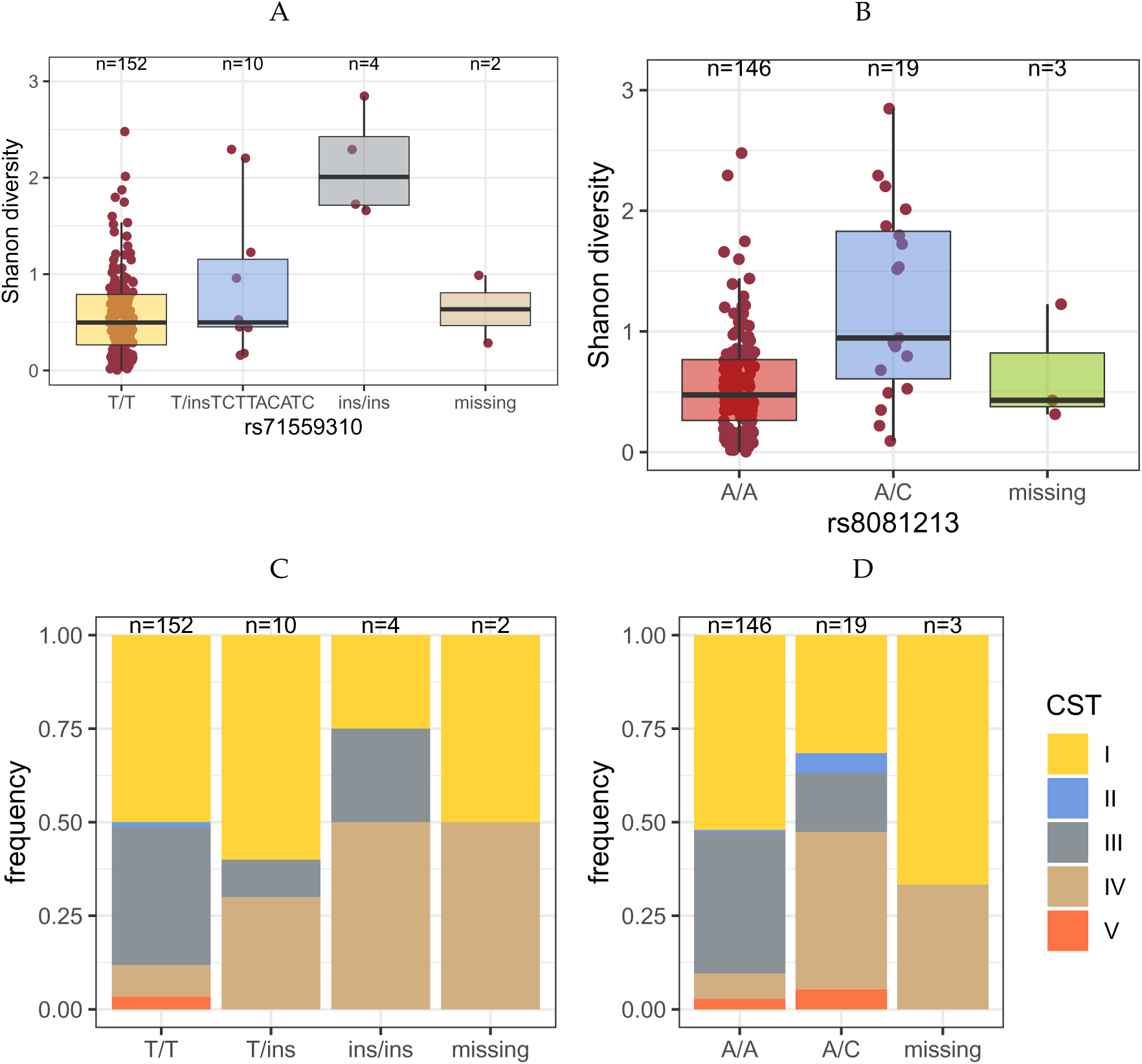
Vaginal microbiota Shannon diversity (A, B) and community state (C, D) type stratified by genetic variant. For some individuals, the coverage for the genomic position is missing. Numbers above the boxes indicate group sample sizes.

**Figure 4:**
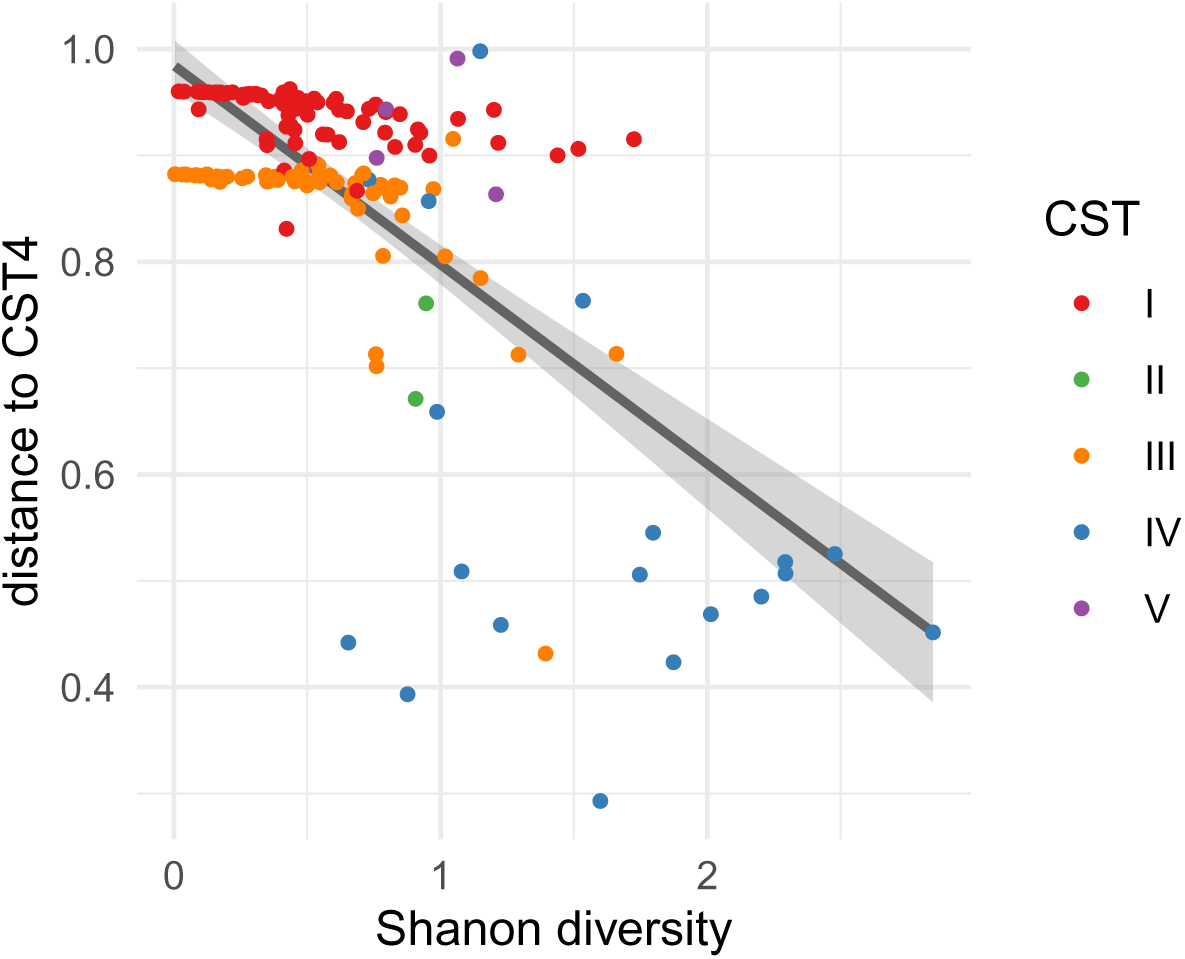
Distribution of the phenotypic traits of interest. Each of the dots corresponds to one of the participants. The line shows the output of the linear model, showing the negative correlation between Shannon diversity and distance to CST4. Colours show the different CSTs (only CST4 is poor in lactobacilli).

### Genetic control over the traits

When considering the explanatory power of all the SNPs on of a trait of interest, the signal was limited when using the binary trait (CST4 *vs.* non-CST4) with a genetic variance of 0.085 (standard error of 0.074) and even more when using the distance to CST4, with a genetic variance of 0.018 (standard error of 0.016). For the Shannon diversity, however, the genetic variance was 0.24 for the whole genome, with a standard error of 0.22. Therefore, we focus on the latter trait in the following.

### Association study

The genome wide association study, assuming an additive model and using the ten major MDS components as covariates, identified two specific genomic regions as being associated with vaginal microbiota Shannon diversity, although not significantly according to the classical 5 *×* 10^−8^ threshold.

The first region we identified on chromosome 8 corresponds to a potential insertion (rs71559310). The second corresponds to a wider region on chromosome 17 with the highest signal being for SNP rs8081231 (Table 2).

**Table 2:** Characteristics of the five SNPs most significantly associated with vaginal microbiota Shannon diversity. CHR stands for chromosome number, POS for position in genome reference GRCh37, REF for the reference allele, ALT for the alternative allele, FREQ for the frequency of the alternative allele, TEST for the test used (here additive), N for the number of samples in the regression, *β* for the regression coefficient, SE for standard error, T for the t-statistic, and P for the asymptotic p-value. For rs71559310, the insertion is TAATACATCATTTCTTACATC.

| CHR | POS | ID | REF | ALT | FREQ | TEST | N | $\beta$ | SE | T | P |
| --- | --- | --- | --- | --- | --- | --- | --- | --- | --- | --- | --- |
| 17 | 25297867 | rs8081213 | A | C | 0.056 | ADD | 161 | 0.71 | 0.12 | 6.02 | 1.32e-08 |
| 17 | 25371993 | rs146879951 | A | G | 0.056 | ADD | 161 | 0.71 | 0.12 | 5.98 | 1.58e-08 |
| 8 | 61775368 | rs71559310 | T | insertion | 0.043 | ADD | 162 | 0.87 | 0.16 | 5.32 | 3.69e-07 |
| 17 | 25344814 | rs25344814 | A | ATT | 0.062 | ADD | 161 | 0.61 | 0.11 | 5.32 | 3.82e-07 |
| 17 | 25294260 | rs73325567 | C | G | 0.061 | ADD | 163 | 0.60 | 0.11 | 5.31 | 3.89e-07 |

We compared the population using these variants as stratification to detect potential divergence in terms of demographic and behavioural covariates. For rs71559310, the only slight difference resided in the percentage of women who have already been pregnant in the past (Table S1), which was higher for women with the insertion associated with CST4; a difference that was already present in the cohort profile (Table 1). For rs8081213, women with the heterozygote genotype (A/C) had a lower age at first menstruation and reported less sports practice (Table S2). There was also a non-significant difference in terms of these women also reporting less condom use by the male partner. In both cases, women with fewer lactobacilli reported less European ancestry.

When performing a linear regression using the Shannon diversity as a response variable, and, as explanatory variables, the variants at the two main genomic positions, the main covariates associated with vaginal microbiota composition (in Tables 1), S1, and S2), as well as the ten first components of the MDS we find that our results are largely unaffected. Besides the two genomic regions we identified, the only variable that is significantly associated with Shannon diversity is the one related to earlier pregnancy (Table 3). Furthermore, the model explains 27 % of the variability in Shannon diversity.

**Table 3:**
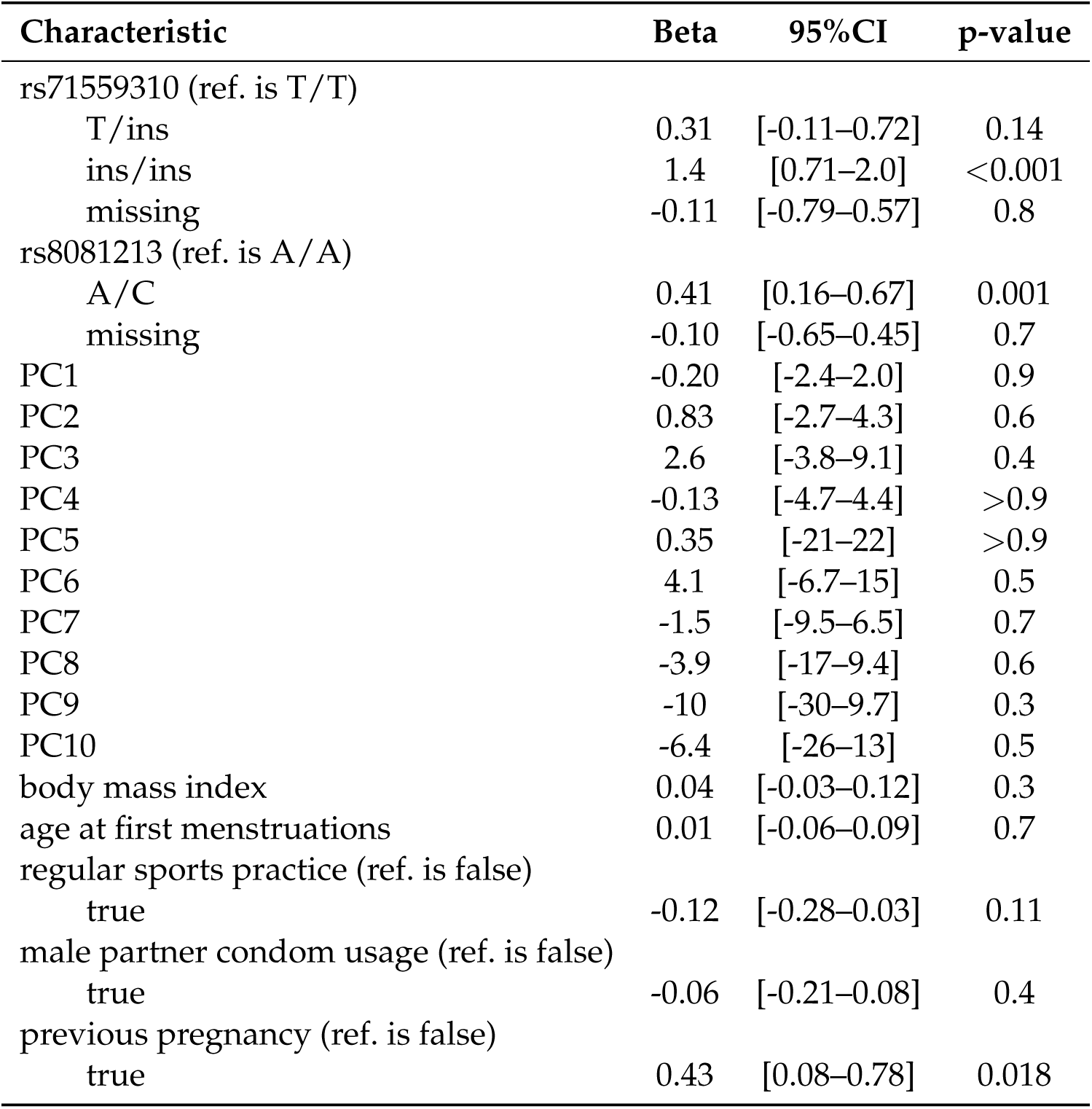
Linear model with socio-demographic covariates. ‘ref.’ indicates the reference value for categorical variables. The adjusted R-squared of the model is 0.271.

## Discussion

Few studies have explored associations between human genetics and the vaginal microbiota. Most of these rely on symptoms rather than genetic microbiological data. Furthermore, few studies have been conducted on women of European ancestry.

Building on data collected during the PAPCLEAR study Kamiya et al. (2025), we performed a GWAS using the Shannon diversity index of the vaginal microbiota as our trait of interest. Our microarray allowed us to identify 320,452 variants, which we enriched to 2,927,125 variants using imputation methods.

Our analysis revealed two associations of interest. First, we found an insertion in chromosome 8 (rs71559310), which is associated with a higher level of Shannon diversity and CST-4. Second, we found a region in chromosome 17, especially one SNP (rs8081231). Both were found on 5.6 % of the chromosomes. This is consistent with the estimated frequencies in human populations of rs8081213, which is 6.9 %. Note also that we did not find any C/C homozygous individuals for this SNP. Interestingly, these variants did not seem to result from differences in demographic and behavioural covariates in the population. Furthermore, together they explained approximately one-fourth of the variance in our trait of interest.

Investigating potential associations with known genes via linkage disequilibrium using tools such as http://locuszoom.sph.umich.edu/ did not yield any significant signal. rs8081231 is far from any coding region. rs71559310 is an intron variant in the gene coding the chromodomain helicase DNA binding protein 7 (CHD7). Pathogenic variants of this gene are associated with the CHARGE syndrome, as well as other pathologies (van Ravenswaaij-Arts et al., 1993).

We made several strong assumptions in this study. The number of observations per participant was highly variable, some being followed for less than two months and others for more than two years (Kamiya et al., 2025). Therefore, we decided to average all the observations of a single participant to define the observed trait. Another possibility could have been to analyse the temporal variability of the vaginal microbiota. However, this would have been problematic for participants with a single sample. More generally, many vaginal microbiota-related treats could have been considered. We focused on Shannon diversity because it was the one for which the genetic variance explained the observed variation best.

Our study is limited in terms of the number of participants, but also of genome coverage. The variants we find do not seem to be correlated with covariates classically involved in vaginal microbiota differences and a deeper sequencing of the human genome could identify positions that were not sufficiently covered in the assay used.

## Methods

### Data generation

189 participants were enrolled in the PAPCLEAR study, which aimed to understand the human papillomavirus kinetics in the vaginal environment (Murall et al., 2019).

DNA was extracted using the QIAamp^®^ DNA mini kit (QIAGEN Inc.) from PBMCs extracted from 10mL of circulating blood following standard protocol for body fluids. Eleven samples were lost during the extraction process, and DNA could not be retrieved. We then analysed the DNA of the 168 participants using an Illumina Global Screening Assay Multi-Disease (GSA-MD) genotyping assay, which targets approximately 800,000 SNPs in the genome.

For each of the participants, we used vaginal microbiota 16S metabarcoding data described in an earlier study (Kamiya et al., 2025).

### Data curation

For the vaginal microbiota data, given the uneven number of samples for each participant, we averaged all the visits to obtain a mean dataset for the relative abundance of 372 bacterial species.

We analysed this dataset for community state type (CST) composition using the VALENCIA nearest centroid classifier (France et al., 2020). From this, we could obtain the distance to each centroid, as well as the CST assignment. We used this data to compute two traits: a binary CST assignment (CST4 *vs.* non-CST4) and a quantitative shortest distance to any CST4.

Using the R diversity function from the vegan package with the mean relative abundance data, we computed our third (quantitative) phenotypic trait, namely the Shannon diversity index.

### Genome Wide Association Study

We performed the association study itself using plink2, using the linear regression with the `--covar` option to use the first ten MDS components as covariates. Only the results assuming an additive genetic effect were kept, and SNPs with p-values higher than 0.01 were ignored for further analyses.

### SNP imputation

In order to enrich our dataset, we imputed genomic positions not directly covered by the GSA-MD assay using IMPUTE2 (Howie et al., 2009). For this, we downloaded the reference genome v.38 and exported each chromosome from the main file using plink v.2.0 (Chang et al., 2015).

### Quality control

We performed classical steps for quality control, as summarised by Marees et al. (2018), namely, we:

- removed variants that were too rare or participants with too many missing variants with a 2 % threshold;
- removed variants with a minority allele frequency with a 5 % threshold;
- checked for a Hardy-Weinberg equilibrium of the variants;
- removed individuals with more than 2 standard deviations in terms of heterozygosity;
- checked for cryptic relatedness, assuming a *π*^threshold value of 0.2.

Using the 1000 Genomes database reference, we also investigated the shared ancestry between our participants. For this, we performed a multidimensional scaling (MDS) clustering, pooling our PAPCLEAR dataset with that from the 1000 Genomes.

The output of this quality control is available in the supplementary HTML file at https://doi.org/10.57745/LGZANK

### Heritability estimation

To help identify the most suitable phenotypic trait for GWAS investigation, we estimated the variance explained by all the SNPs using GCTA (Yang et al., 2011).

### Data availability

The raw genotyping data cannot be made publicly available for ethical reasons. However, the output of the GWAS for Shannon diversity, the metadata, and the output of the quality control analysis can be accessed at https://doi.org/10.57745/LGZANK, along with the scripts used to generate the figures and table in the manuscript.

## Ethics

The PAPCLEAR study has been approved by the Comité de Protection des Personnes (CPP) Sud Méditerranée I (reference number 2016-A00712-49); by the Comité Consultatif sur le Traitement de l’Information en matière de Recherche dans le domaine de la Santé (reference number 16.504); by the Commission Nationale Informatique et Libertés (reference number MMS/ABD/AR1612278, decision number DR-2016-488), by the Agence Nationale de Sécurité du Médicament et des Produits de Santé (reference 20160072000007), and is registered at ClinicalTrials.gov under the ID NCT02946346.

## Acknowledgements

The authors acknowledge the ISO 9001 certified IRD i-Trop HPC (member of the South Green Platform) at IRD Montpellier for providing HPC resources that have contributed to the research results reported within this article (bioinfo.ird.fr and www.southgreen.fr).

## S1 Authors’ contribution

Samuel Alizon conceived the study, performed the statistical analyses, generated the figures, and wrote he initial version of the manuscript.

Claire Bernat, Vanina Boué, Sophie Grasset, Soraya Groc, and Massilva Rahmoun generated the clinical data.

Tsuksuhi Kamiya, Christian Selinger, and Nicolas Tessandier structured the microbiota data and the metadata.

Jacques Ravel and his team generated the vaginal microbiota data.

Jacques Reynes, Michel Segondy, Vincent Foulongne, Marine Bonneau, Christelle Graf, Vincent Tribout, Nathalie Boulle, and Carmen Lia Murall contributed to the conception of the study and supervised the acquisition of the data.

Vincent Pedergnana provided critical input for the statistical analyses. Jean-Franrois Deleuze and his team generated the human genetics data. All authors contributed to the final version of the manuscript.

## S2 Supplementary Results

**Table S1:** Cohort profile stratified by mutation at rs71559310. Only the significant variables are shown. See Table 1 for details.

|  | T/T | T/ins | ins/ins | missing | p-value |
| --- | --- | --- | --- | --- | --- |
| n | 152 | 10 | 4 | 2 |  |
| previous pregnancy (%) | 5 (3.3) | 2 (20.0) | 0 (0.0) | 1 (50.0) | 0.017 |
| caucasian (%) | 133 (87.5) | 2 (20.0) | 1 (25.0) | 0 (0.0) | <0.001 |
| Lactobacillus-CST (%) | 140 (92.1) | 7 (70.0) | 2 (50.0) | 1 (50.0) | 0.004 |
| Shanon diversity | 0.50 [0.26, 0.79] | 0.50 [0.45, 1.16] | 2.01 [1.71, 2.43] | 0.64 [0.46, 0.81] | 0.008 |

**Table S2:** Cohort profile stratified by mutation at rs8081213. Only the significant variables at a 10% threshold are shown. See Table 1 for details.

|  | A/A | A/C | missing | p |
| --- | --- | --- | --- | --- |
| n | 146 | 19 | 3 |  |
| regular sport practice (%) | 78 (53.4) | 6 (31.6) | 0 (0.0) | 0.032 |
| male partner condom usage (%) | 81 (55.5) | 6 (31.6) | 1 (33.3) | 0.074 |
| age at first menstruations | 0.19 [-0.55, 0.93] | -0.55 [-0.55, 0.19] | -1.30 [-1.67, -0.93] | 0.055 |
| identified as caucasian (%) | 123 (84.2) | 12 (63.2) | 1 (33.3) | 0.013 |
| Lactobacillus-dominated (%) | 137 (93.8) | 11 (57.9) | 2 (66.7) | <0.001 |
| Shannon diversity | 0.48 [0.26, 0.77] | 0.95 [0.60, 1.84] | 0.43 [0.37, 0.83] | 0.001 |

## References

Bilardi JE, Walker S, Temple-Smith M, McNair R, Mooney-Somers J, Bellhouse C, et al. 2013. The burden of bacterial vaginosis: women’s experience of the physical, emotional, sexual and social impact of living with recurrent bacterial vaginosis. PLOS ONE 8:e74378. doi:10.1371/journal.pone.0074378.

Borgdorff H, Veer Cvd, Houdt Rv, Alberts CJ, Vries HJd, Bruisten SM, et al. 2017. The association between ethnicity and vaginal microbiota composition in Amsterdam, the Netherlands. PLOS ONE 12:e0181135. doi:10.1371/journal.pone.0181135.

Brotman RM, Ghanem KG, Klebanoff MA, Taha TE, Scharfstein DO, Zenilman JM. 2008. The effect of vaginal douching cessation on bacterial vaginosis: a pilot study. American Journal of Obstetrics and Gynecology 198:628.e1–628.e7. doi:10.1016/j.ajog.2007.11.043.

Chang CC, Chow CC, Tellier LC, Vattikuti S, Purcell SM, Lee JJ. 2015. Second-generation PLINK: rising to the challenge of larger and richer datasets. GigaScience 4:s13742–015–0047–8. doi:10.1186/s13742-015-0047-8.

Fan W, Kan H, Liu HY, Wang TL, He YN, Zhang M, et al. 2021. Association between Human Genetic Variants and the Vaginal Bacteriome of Pregnant Women. mSystems 6:10.1128/msystems.00158-21. doi:10.1128/msystems.00158-21.

Fettweis JM, Brooks JP, Serrano MG, Sheth NU, Girerd PH, Edwards DJ, et al. 2014. Differences in vaginal microbiome in african american women versus women of european ancestry. Microbiology 160:2272.

France MT, Ma B, Gajer P, Brown S, Humphrys MS, Holm JB, et al. 2020. VALENCIA: a nearest centroid classification method for vaginal microbial communities based on composition. Microbiome 8:166. doi:10.1186/s40168-020-00934-6.

Haahr T, Zacho J, Bräuner M, Shathmigha K, Skov Jensen J, Humaidan P. 2019. Reproductive outcome of patients undergoing in vitro fertilisation treatment and diagnosed with bacterial vaginosis or abnormal vaginal microbiota: a systematic PRISMA review and meta-analysis. BJOG 126:200–207. doi:10.1111/1471-0528.15178.

Howie BN, Donnelly P, Marchini J. 2009. A Flexible and Accurate Genotype Imputation Method for the Next Generation of Genome-Wide Association Studies. PLOS Genetics 5:e1000529. doi:10.1371/journal.pgen.1000529.

Kamiya T, Tessandier N, Elie B, Bernat C, Boué V, Grasset S, et al. 2025. Factors shaping vaginal microbiota long-term community dynamics in young adult women. Peer Community Journal 5:e30. doi:10.24072/pcjournal.527.

Kurilshikov A, Medina-Gomez C, Bacigalupe R, Radjabzadeh D, Wang J, Demirkan A, et al. 2021. Large-scale association analyses identify host factors influencing human gut microbiome composition. Nature Genetics 53:156–165. doi:10.1038/s41588-020-00763-1.

Ma B, Forney LJ, Ravel J. 2012. Vaginal Microbiome: Rethinking Health and Disease. Annual Review of Microbiology 66:371–389. doi:10.1146/annurev-micro-092611-150157.

Marees AT, de Kluiver H, Stringer S, Vorspan F, Curis E, Marie-Claire C, et al. 2018. A tutorial on conducting genome-wide association studies: Quality control and statistical analysis. International Journal of Methods in Psychiatric Research 27:e1608. doi:10.1002/mpr.1608.

McKinnon LR, Achilles SL, Bradshaw CS, Burgener A, Crucitti T, Fredricks DN, et al. 2019. The Evolving Facets of Bacterial Vaginosis: Implications for HIV Transmission. AIDS Research and Human Retroviruses 35:219–228. doi:10.1089/aid.2018.0304.

Mehta SD, Nannini DR, Otieno F, Green SJ, Agingu W, Landay A, et al. 2020. Host Genetic Factors Associated with Vaginal Microbiome Composition in Kenyan Women. mSystems 5:10.1128/msystems.00502–20. doi:10.1128/msystems.00502-20.

Murall CL, Rahmoun M, Selinger C, Baldellou M, Bernat C, Bonneau M, et al. 2019. Natural history, dynamics, and ecology of human papillomaviruses in genital infections of young women: protocol of the PAPCLEAR cohort study. BMJ Open 9:e025129. doi: 10.1136/bmjopen-2018-025129.

Murphy K, Shi Q, Hoover DR, Adimora AA, Alcaide ML, Brockmann S, et al. 2024. Genetic predictors for bacterial vaginosis in women living with and at risk for HIV infection. American Journal of Reproductive Immunology 91:e13845. doi:10.1111/aji.13845.

Mutli E, Mändar R, Koort K, Salumets A, Team EBR, Laisk T. 2024. Genome-wide association study in Estonia reveals importance of vaginal epithelium associated genes in case of recurrent vaginitis. Journal of Reproductive Immunology 162:104216. doi: 10.1016/j.jri.2024.104216.

Ravel J, Gajer P, Abdo Z, Schneider GM, Koenig SSK, McCulle SL, et al. 2011. Vaginal microbiome of reproductive-age women. Proc Nat Acad Sci USA 108:4680–4687. doi: 10.1073/pnas.1002611107.

Tamarelle J, Thiébaut ACM, de Barbeyrac B, Bébéar C, Bourret A, Fauconnier A, et al. 2024. Vaginal microbiota stability over 18 months in young student women in France. European Journal of Clinical Microbiology & Infectious Diseases 43:2277–2292. doi:10.1007/s10096-024-04943-3.

Tessandier N, Elie B, Boué V, Selinger C, Rahmoun M, Bernat C, et al. 2025. Viral and immune dynamics of genital human papillomavirus infections in young women with high temporal resolution. PLOS Biology 23:e3002949. doi:10.1371/journal.pbio.3002949.

van de Wijgert JHHM. 2017. The vaginal microbiome and sexually transmitted infections are interlinked: Consequences for treatment and prevention. PLOS Medicine 14:e1002478. doi:10.1371/journal.pmed.1002478.

van Ravenswaaij-Arts CM, Hefner M, Blake K, Martin DM. 1993. CHD7 Disorder. In: Adam MP, Feldman J, Mirzaa GM, Pagon RA, Wallace SE, Amemiya A, eds., GeneReviews®, Seattle (WA): University of Washington, Seattle. URL http://www.ncbi.nlm.nih.gov/books/NBK1117/.

Yang J, Lee SH, Goddard ME, Visscher PM. 2011. GCTA: A Tool for Genome-wide Complex Trait Analysis. The American Journal of Human Genetics 88:76–82. doi: 10.1016/j.ajhg.2010.11.011.

Zhou X, Brown CJ, Abdo Z, Davis CC, Hansmann MA, Joyce P, et al. 2007. Differences in the composition of vaginal microbial communities found in healthy Caucasian and black women. ISME J 1:121–133. doi:10.1038/ismej.2007.12.

